# Understanding Uncertainty in Large Language Model Predictions of Early Death in Critically Ill Patients: A Conformal Prediction Approach

**DOI:** 10.64898/2025.12.14.25342221

**Authors:** Fatemeh Shah-Mohammadi, Alexander Millar, Julio Facelli, Ramkiran Gouripeddi

## Abstract

**Background:** Early prediction of in-hospital death remains a significant challenge due to the limited availability of structured data during initial admission. Unstructured clinical notes, which often contain important observations and impressions, are an underutilized resource for real-time risk stratification. While leveraging recent advances in large language models (LLM) is a promising approach to use this unstructured information, the lack of understanding of the uncertainty of LLM predictions, at the patient level, for such critical forecasts is a serious deterrence for their use in clinical settings.

**Objective:** This study aims to evaluate the effectiveness and confidence, in predicting in-hospital death probability for an individual patient using LLMs, specifically GPT-4o and unstructured clinical notes.

**Methods:** We applied conformal prediction to quantify the uncertainty of GPT-4o’s zero-shot predictions for in-hospital death, leveraging concatenated clinical notes documented from the first 24 hours of intensive care unit (ICU) admission in MIMIC-III for patients with acute kidney failure who were admitted through the emergency department (ED).

**Results:** Across both classes “in-hospital death” and “in-hospital survive”, the GPT model performed better on the in-hospital death class, achieving precision 0.52 (95% CI 0.48–0.56), recall 0.93 (95% CI 0.90–0.95), and F1-score 0.66 (95% CI 0.63– 0.70). The conformal prediction (CP) framework provided an overall empirical coverage of 90.4%, exceeding the target threshold of 90%. However, class-specific coverage was imbalanced, with 99.7% for the death and 81.1% for the survived class.

**Conclusions:** The model’s outputs exhibit overconfidence, particularly in cases of incorrect predictions. Integrating conformal prediction provides a promising approach to quantifying and calibrating uncertainty in large language model outputs for individual patient predictions, thereby enhancing their potential applicability for clinical decision-making.

## Introduction

In the early hours following emergency admission, resource constraints and sparse structured data make continuous risk assessment difficult, particularly for patients at risk of life-threatening organ dysfunction or unexpected death [1–4]. Predicting death among critically ill patients is also valuable for evaluating the effectiveness of treatments, clinical guidelines, surgeries, and other interventions using electronic health record (EHR) data [5]. These records typically include demographic details, physiological data, early lab results, clinical observations, and treatment information [6,7]. Various scoring systems, such as the Emergency Severity Index [8], APACHE (Acute Physiology and Chronic Health Evaluation) [9], Death Probability Models [10], and the Sequential Organ Failure Assessment (SOFA) score [11]—have been developed to assess illness severity and predict patient outcomes. However, despite their widespread use, these models often have limited accuracy and are primarily used as benchmarking tools [12,13]. Moreover, the relative accuracy varies substantially among patients and methods to quantify prediction reliability at the patient level remain poorly established.

Machine learning (ML) promised better performance by leveraging electronic health records (EHRs) and publicly available databases such as medical information mart for intensive care (MIMIC-III) [14] and high time resolution ICU dataset (HiRID) [15], and community challenges have showcased ML’s ability to learn complex, nonlinear relations among physiological variables [16,17] and predict patient outcomes, including ICU death. However, many ML pipelines rely on curated, structured input features and assume stable data availability, conditions often unmet during early stages of patient care [48,54]. Moreover, ML systems typically fail to provide clinicians with calibrated, case-level uncertainty estimates, making it difficult to determine when predictions are trustworthy [49, 55]. Large language models (LLMs) change this equation by circumventing restrictive data assumptions and directly consuming unstructured clinical notes, often the richest source of information available in the first 24 hours, while requiring minimal task-specific training [50–53]. Unlike traditional ML, LLMs can capture nuanced clinical context through advanced language comprehension [18–24,56–57]. Models like Generative Pre-trained Transformer (GPT) have shown promise in this space, performing at or above human level on standardized medical licensing exams and demonstrating the ability to extract relevant insights from complex clinical documentation [21]. Their success is largely attributed to their vast pretraining data, scale of parameters, and fine-tuning through reinforcement learning with human feedback (RLHF) [22], making them a compelling addition to predictive modeling in healthcare [23–33,50]. Building on this, authors in [56] evaluated GPT for real-world ED acuity assessment and found that a general-purpose model could approximate clinician triage decisions, while authors in [57] assessed GPT in scenario-based ED triage tasks and reported feasible but variable performance. These studies underscore the growing applicability of LLMs in emergency care settings, where rapid, text-based clinical reasoning can augment early decision support. Extending this direction, our study leverages LLMs to predict in-hospital death based solely on unstructured clinical notes documented within the first 24 hours of an ICU patient’s admission, aiming to enable early risk stratification and clinical prioritization in critically ill patients admitted through the ED. Prior work has explored death prediction using structured data and text-derived variables from the first 48 hours of hospitalization [51] and compared performance of few traditional ML models, with and without unstructured text, and reported improved discrimination when including manually engineered text features. Another study [52] relied exclusively on notes and traditional ML pipelines to predict short-, mid-, and long-term ICU death, using feature extraction methods like frequency-based and embedding-based representations. In contrast, our work evaluates a general-purpose, non-fine-tuned LLM model operating in a zero-shot setting, consuming raw clinical narratives without any handcrafted preprocessing or retraining. Moreover, since LLMs are known to occasionally produce information that is not factually accurate or grounded in reality, commonly referred to as “hallucinations” [34], quantitatively assessing the reliability of their outputs at the patient level is critical. To responsibly support clinical decision-making, we must quantify the uncertainty associated with LLM predictions at the patient level [55]. To quantify this uncertainty, we propose to adopt the conformal prediction (CP) framework, a model-agnostic, distribution-free approach that offers rigorous coverage guarantees [35–37]. CP allows to set a desired error rate (α) and generates prediction sets that contain the true label with a guaranteed level of confidence. By using nonconformity scores—such as 1-*f(X)_Y_*, where *f(X)_Y_* is the softmax score for the true label [38] CP provides a practical and interpretable method to express and control the uncertainty in LLM-generated predictions, which is essential for supporting trustworthy clinical decision-making. Using nonconformity scores, CP yields interpretable error control at the patient level, directly addressing the gap left by traditional scores and standard ML.

Building on this rationale, we investigate whether a LLM (GPT-4o) can predict in-hospital death from clinical notes recorded within the first 24 hours of admission and, critically, whether CP can quantify and communicate prediction uncertainty in a clinically meaningful way. Unlike prior work that relies on structured data or lacks calibrated uncertainty, our approach operates directly on unstructured intensive care unit (ICU) clinical notes from the first 24 hours of care for patients admitted through ED, providing explicit, controllable error guarantees via conformal prediction. To our knowledge, no prior study has jointly evaluated GPT-4o for this task and paired it with a CP framework to provide calibrated, patient-level uncertainty on unstructured notes.

## Methods

### Dataset

The dataset used in this study is MIMIC-III, a widely recognized and comprehensive repository of de-identified health data [14]. It encompasses clinical information from approximately 60,000 intensive care unit patients treated at Beth Israel Deaconess Medical Center in Boston, Massachusetts, across nearly a decade. This dataset includes a wide range of information, such as electronic health records, laboratory results, medication prescriptions, and clinical notes. As a first step to demonstrate the application of our methods, we focused on patients who were initially admitted through the emergency department, with an admission type labeled as “Emergency”, admission location recorded as “Emergency Room Admit” and an initial diagnosis of Acute kidney failure (584.9). We centered our study on patients diagnosed with acute kidney failure because this condition is both clinically significant and prevalent among critically ill patients, often serving as a marker of multi-organ dysfunction and poor prognosis. Early detection of deterioration in this population is particularly valuable, as timely interventions such as renal replacement therapy or hemodynamic optimization can meaningfully alter outcomes. Moreover, acute kidney failure’s heterogeneous presentation in unstructured clinical notes provides a rigorous and challenging test case for evaluating language models’ ability to extract prognostic signals— and, through conformal prediction, to quantify uncertainty at the patient level— from narrative documentation within the first 24 hours of ICU care (following ED admission). For these patients, we retrieved the clinical notes documented within the first 24 hours of ICU patient admission. All available notes from this period were concatenated and submitted to the LLM. For consistency in the following sections, we will address this concatenated set of clinical notes as the “clinical note”. The final study cohort consisted of 4,714 unique admissions, among whom 910 patients (19.3%) died during their hospital stay forming the **“***in-hospital death”* class, while 3,804 patients (80.7%) survived forming the **“***in-hospital survived” class*. To address the class imbalance in the dataset, we under-sampled the patients who survived to create a balanced cohort. After removing the outliers, the final cohort consisted of 1,354 records. To maintain balance in the calibration and test sets for conformal prediction, we performed a stratified split followed by majority class under-sampling where necessary.

### Conformal prediction

Most current CP methods for LLMs depend on access to model logits to compute nonconformity scores. Model logits are the raw, unnormalized output scores that reflect the model’s confidence in each possible token and are produced by the model before applying the softmax function to obtain probabilities. Authors in [39] defined nonconformity scores using softmax outputs over logits for multiple-choice question answering (MCQ) tasks, while references [40,41] apply the conformal risk control framework, a generalization of CP, to LLMs by leveraging log-probabilities derived from the model. However, for API-only LLMs, end users typically do not have access to model logits. Even when logits are available, they are often miss calibrated, which can impair the effectiveness of CP by yielding unreliable prediction sets or intervals, often resulting in large set sizes and thus reduced efficiency [42–44]. The framework introduced in [45] offers a complementary approach by avoiding logit dependence altogether and instead quantifies uncertainty using entropy calculated from model-assigned confidence scores. This entropy-based method enabled staged decision-making with selective human oversight.

In our work, we implemented the framework introduced in [44], with a slight modification. To implement conformal prediction in the absence of direct access to model logits, we initially sampled 30 [44] randomly independent responses per each patient input and used the empirical frequency of each response as a coarse-grained measure of uncertainty. The sampling rate of 30 responses per input was selected following prior work [44], which acknowledges that while thousands of samples are theoretically required to approximate true predictive probabilities with high precision, such exhaustive sampling is computationally not feasible. As a result, a smaller, tractable number, such as 30, offers a pragmatic balance between uncertainty estimation and computational efficiency. For each prediction, the LLM reports one answer, either “Yes” or “No”, but this frequency-based approach introduces a key limitation: multiple responses may share the same frequency count despite differing in underlying uncertainty levels [44]. This results in a concentration of nonconformity scores on a limited set of values, thereby reducing the efficiency of the resulting prediction sets. To address this limitation, we enhanced the approach by capturing the top 10 log-probabilities generated for each sample.

Figure 1 shows an overview of the proposed conformal prediction pipeline, divided into three main stages: Prompting & Sampling, Post-Processing, and Nonconformity Score. In the first stage, concatenated clinical notes documenting the first 24 hours of admission per patient are combined with a structured prompt and submitted to GPT-4o. From this point onward, any reference to a “note” refers to this concatenated set of notes from the first 24 hours of admission for each patient. To approximate uncertainty without access to internal logits, the model is queried 30 times receiving 30 randomly independent responses per patient *i* input. Each response is stored together with the top-K=10 candidate tokens {t*_ir_*1*t*, … {_*irK*_} predicted at the response position and their associated log-probabilities {_*ir*1_, … {*_irK_*}. We convert and (slightly) renormalize the probabilities. The cumulative sum of top 10 probabilities consistently reached a threshold of 0.99 that enables uniform and reliable normalization across all samples.

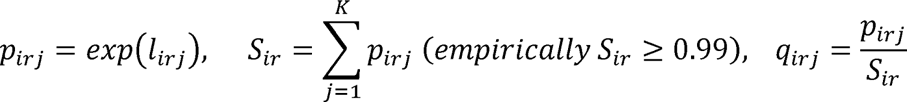

**Figure 1:**
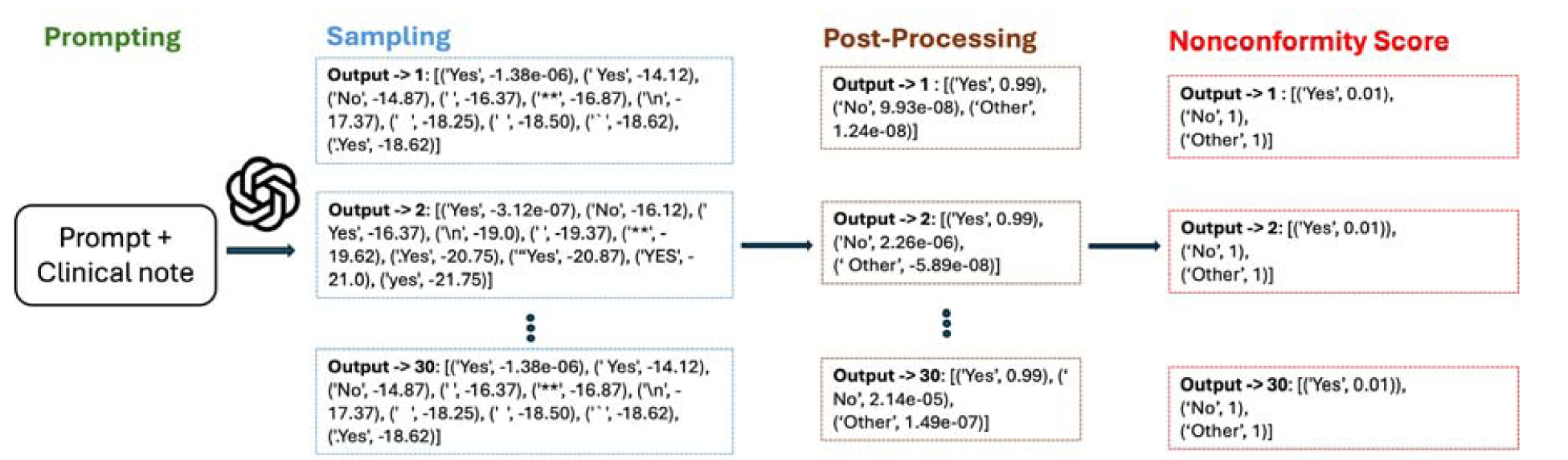
Overview of the conformal prediction pipeline. (Prompting & Sampling) Concatenated ICU notes from the first 24 hours of admission are prompted to GPT-4o. Each note is queried 30 times; for every response, the top 10 tokens and log-probabilities are captured, exponentiated, and normalized. (Post-Processing): Tokens are lower-cased, mapped to classes via indicator functions, and aggregated class probabilities are calculated for each response. (Nonconformity Score): For each response, class-specific nonconformity scores are computed.

To ensure consistency during post-processing, we normalized the tokens by converting all candidates to lowercase. In the post-processing stage, probabilities are converted into class-level probabilities by mapping each token to a class using indicator functions:

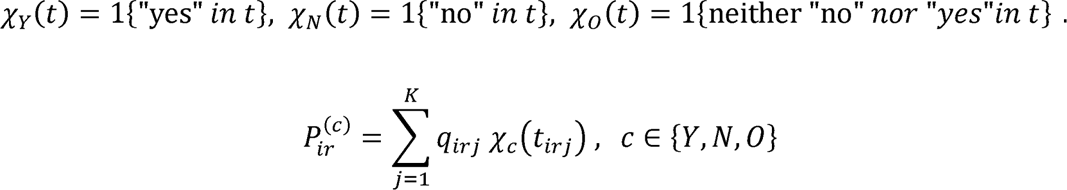

Specifically, we computed the total probability mass assigned to responses containing the word “yes” as the probability of the Y= “*in-hospital death*” class, and similarly aggregated probabilities mass assigned to responses containing the word “no” for the N= “*in-hospital survived*” class. Probabilities for responses not containing either keyword have been aggregated and, for illustration purposes in Figure 1, are represented under a class named O= “other.” It is worth noting that we have only two main classes in this task: “in-hospital survived,” illustrated as “No” in this figure, and “in-hospital death,” illustrated as “Yes.” The high probabilities are concentrated within the top 10 tokens, cumulatively summing to approximately 0.99, while the remaining token probabilities are negligibly small. This confirms that the probability mass is effectively captured within the top 10 candidates. The one-word point label for that query is:

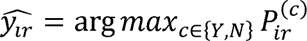

In the final stage, nonconformity scores 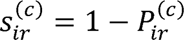 are computed for each prediction. This method provided a more fine-grained and probabilistically grounded estimate of uncertainty while remaining compatible with API-based LLMs.

### LLM Methods

In this study we used the GPT-4o model for generative question answering [46]. We accessed this model through the chat completions API endpoint in a zero-shot setting. In zero-shot prediction, a model is presented with tasks or queries for which it has not received explicit training. It is expected to extrapolate knowledge from its pre-existing understanding of language and context to generate meaningful responses.

Since prompt engineering is essential when interacting with any LLM to obtain high-quality responses, we first experimented with and formulated prompts to elicit the desired responses from the model. The examined prompts are provided in Appendix 1. Then we used our finalized prompt in a zero-shot prediction setting. Our finalized prompt was selected to be as follows:

> “You are a clinical risk prediction model. I will provide you with a clinical note recorded on the first day of a patient’s hospital admission. Based solely on the information provided in the note, your task is to predict whether the patient is at high risk of in-hospital death during their current admission. Please carefully analyze the content of the note for clinical indicators such as vital signs, symptoms, laboratory results, and any other documented clinical findings that might signal a critical condition. Your response must be exactly one word: “Yes” if the note suggests that the patient is at risk of in-hospital death, or “No” if it does not. Do not include any additional text or explanation: <clinical note text here>”.Those clinical notes for which the LLM answers “Yes” will be associated to the “in-hospital death” class, while those receiving a “No” answer will be associated with **“**in-hospital survived” class. To align with emerging standards for transparency and reproducibility in LLM research, we followed the checklist introduced in [47], which provides structured guidance for reporting applied studies using pretrained models with minimal modifications.

Evaluation was conducted on a balanced dataset, and calibration was achieved using an α of 0.1 to ensure 90% coverage. The calibration set was constructed by randomly selecting 50% of the cohort, with the remaining 50% used for testing.

## Results

For the LLM-only (pre-CP) evaluation, aggregating performance across both classes (in-hospital death and survival), the GPT model achieved an overall accuracy of 53%, precision of 58%, recall of 53%, and an F1 score of 44%. When stratified by class, recall was 93% for in-hospital death and 13% for in-hospital survival, showing imbalance in class-specific sensitivity. This observed asymmetry indicates that the model preferentially flags high-risk (death) cases, an operating point that can be beneficial for early warning in high-risk critical-care settings (as in our cohort of ED-admitted patients with acute kidney failure requiring ICU care)

To assess potential demographic biases in our dataset, we examined the distribution of in-hospital death across ethnicity, gender, and marital status, as summarized in Appendix 2.

The distribution of token counts per patient note exhibited substantial variability across the cohort. Overall, the average number of tokens per patient was 1,321 for those who died during hospitalization and 1,707 for those who survived. The standard deviations were also large, 909 and 2,022 respectively, indicating a wide spread in note lengths. Notably, patients who survived had longer clinical notes on average, suggesting that survival cases may involve more extensive documentation.

When examining classification performance by subgroup, correctly classified death cases had a mean token count of 1,376 (SD: 906), while misclassified ones had significantly fewer tokens (mean: 461, SD: 362). An independent samples t-test confirmed the significance of this difference (t = 6.35, p = 3.88 × 10^-10^), suggesting that longer notes are associated with more accurate death predictions, likely due to the richer clinical detail they provide. In contrast, among patients who survived, correctly classified cases had a mean of 922 tokens (SD: 1,537), whereas misclassified survival cases, those incorrectly predicted, had substantially longer notes (mean: 1,812, SD: 2,057). This difference was also statistically significant (t = -3.76, p = 1.87 × 10^-5^), with the negative t-statistic indicating that excessive documentation may contribute to false positives. These findings suggest that while longer notes generally aid prediction in death cases, they may introduce ambiguity in survival cases. To probe this ambiguity, we conducted a content-focused review of both short and long notes across survival cases and compared correctly and incorrectly classified examples. Among short notes of comparable length, correctly predicted survivors emphasized a coherent recovery trajectory (e.g., weaning off vasopressors, improved oxygenation, stable hemodynamics), whereas misclassified survivors contained mixed or competing severity cues (persistent hypoxemia/wheezing, rising creatinine, tachyarrhythmia, marked hyperglycemia) that likely elevated perceived risk. Extending this analysis to long survival notes misclassified as death cases, we observed dense clusters of high-acuity terminology (e.g., sepsis, aspiration/pneumonia/atelectasis, massive hemoptysis), intervention-heavy narratives (intubation, paralytics, vasopressors, blood products), and repeated radiology blocks with hedged differentials (“versus,” “could reflect,” “cannot be excluded”). These features amplify perceived risk, particularly when temporal resolution is unclear and improvement cues are sparse, transient, or embedded within high-acuity context; comorbidity priors (e.g., metastatic disease) and copy-forward repetition further magnify severity signals, while negated or qualified improvements are linguistically weaker and appear underweighted by the model. Collectively, these observations suggest that semantic coherence and a resolved clinical trajectory, rather than length per se, are the principal determinants of correct classification; notes that clearly establish either improvement or deterioration are classified accurately regardless of length, whereas those mixing competing severity cues or lacking temporal resolution—particularly when lengthy—are prone to misclassification.

For the conformal prediction framework, an alpha value of 0.1 was chosen, targeting 90% coverage. This calibration distribution is used to determine the nonconformity threshold that ensures prediction sets on unseen data will include the true label with at least 90% probability.

Figure 2 presents the histogram and density plots of nonconformity scores derived from GPT-4o responses on the calibration set. Each nonconformity score reflects how uncertain the model was about the correct label, computed as 1-*p*(True class), based on the class probability derived from the top 10 log-probabilities across 30 sampled responses per patient. The plot on the left shows the distribution of scores for patients who survived (*in-hospital survived* class), while the right plot shows the scores for patients who died (*in-hospital death* class). In the case of patients who died, the majority of nonconformity scores are clustered near zero. This indicates that the model assigned high confidence (i.e., high probability) to the correct class label. Conversely, for patients who survived, nonconformity scores are generally higher, with a notable shift toward 1.0. This shows that the model often assigned low probability to the correct survival label, implying higher uncertainty or overconfidence in incorrect predictions.

**Figure 2:**
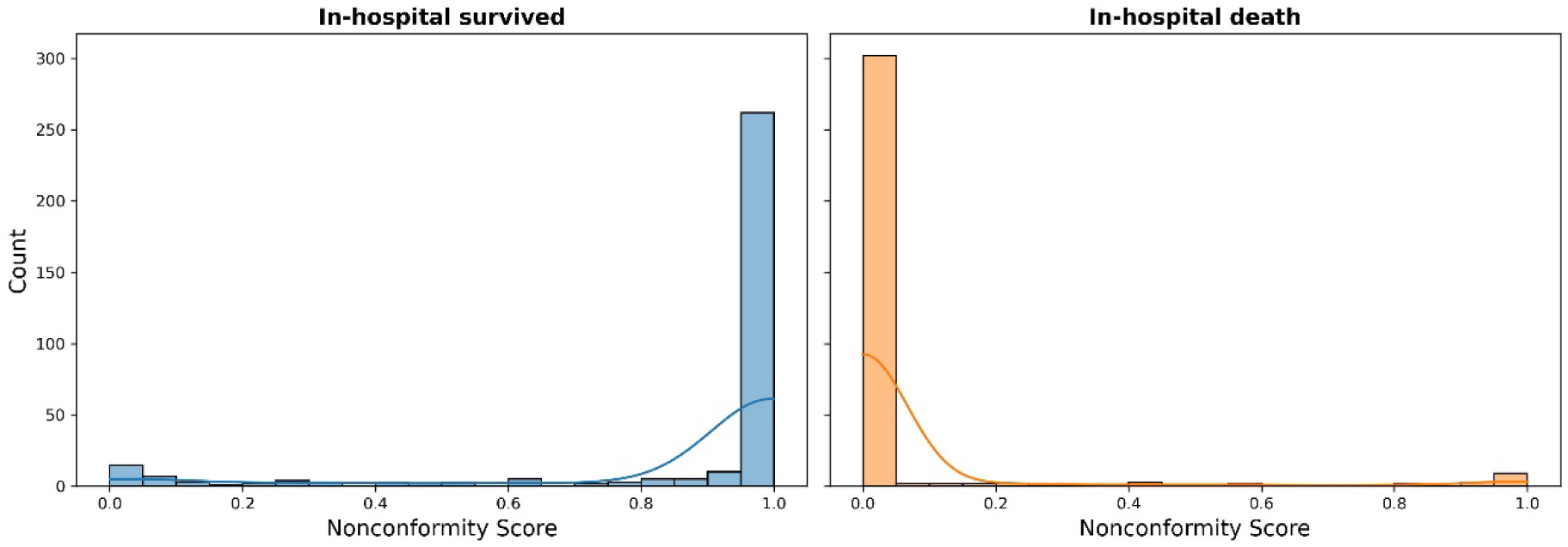
Distribution of nonconformity scores by outcome class in the calibration set, with blue indicating survival and orange indicating in-hospital death.

Figure 3 shows the relationship between the length of clinical notes (in tokens, y-axis) and the corresponding nonconformity scores (x-axis) for each patient in the calibration set, stratified by in-hospital death outcome. The distribution of note categories recorded within the first 24 hours of hospital admission is as follow: “Radiology” (40.48%) and “Nursing/other” (31.97%) categories dominate the dataset and collectively accounting for over 70% of all early documentation. This highlights the intensive use of diagnostic imaging and nursing observations in the immediate phase of patient assessment and care. The next most common category of notes are “Nursing” (14.36%) and “Physician” (10.46%). Categories “Nursing” and “Nursing/other” capture vitals, observations and symptom tracking while physician notes contain diagnostic and treatment plans. The remaining note categories, including “General”, “Respiratory”, “Case Management”, “Consults”, “Nutrition”, “Pharmacy”, “Rehab Services”, and “Social Work”, make up a small fraction of the early documentation. This reflects their limited involvement during the initial 24-hour window, which is typically focused on acute medical assessment and stabilization. Consistent with previous findings, the plot reveals that nonconformity scores for patients who died tend to be low across a range of note lengths, indicating that the model often assigns high probability to the correct death prediction in these cases. In contrast, surviving patients exhibit a broader spread of nonconformity scores, with many clustered near 1.0, especially at the extreme ends of the note length spectrum. Notably, a dense cluster of long survival notes (far right) is associated with high nonconformity scores, suggesting that the model becomes overconfident in incorrect death predictions when presented with excessively lengthy documentation. Overall, this figure reinforces the insight that both insufficient and excessively detailed notes contribute to model uncertainty and misclassification, particularly for survival cases which are aligned with data presented in Table 1.

**Figure 3:**
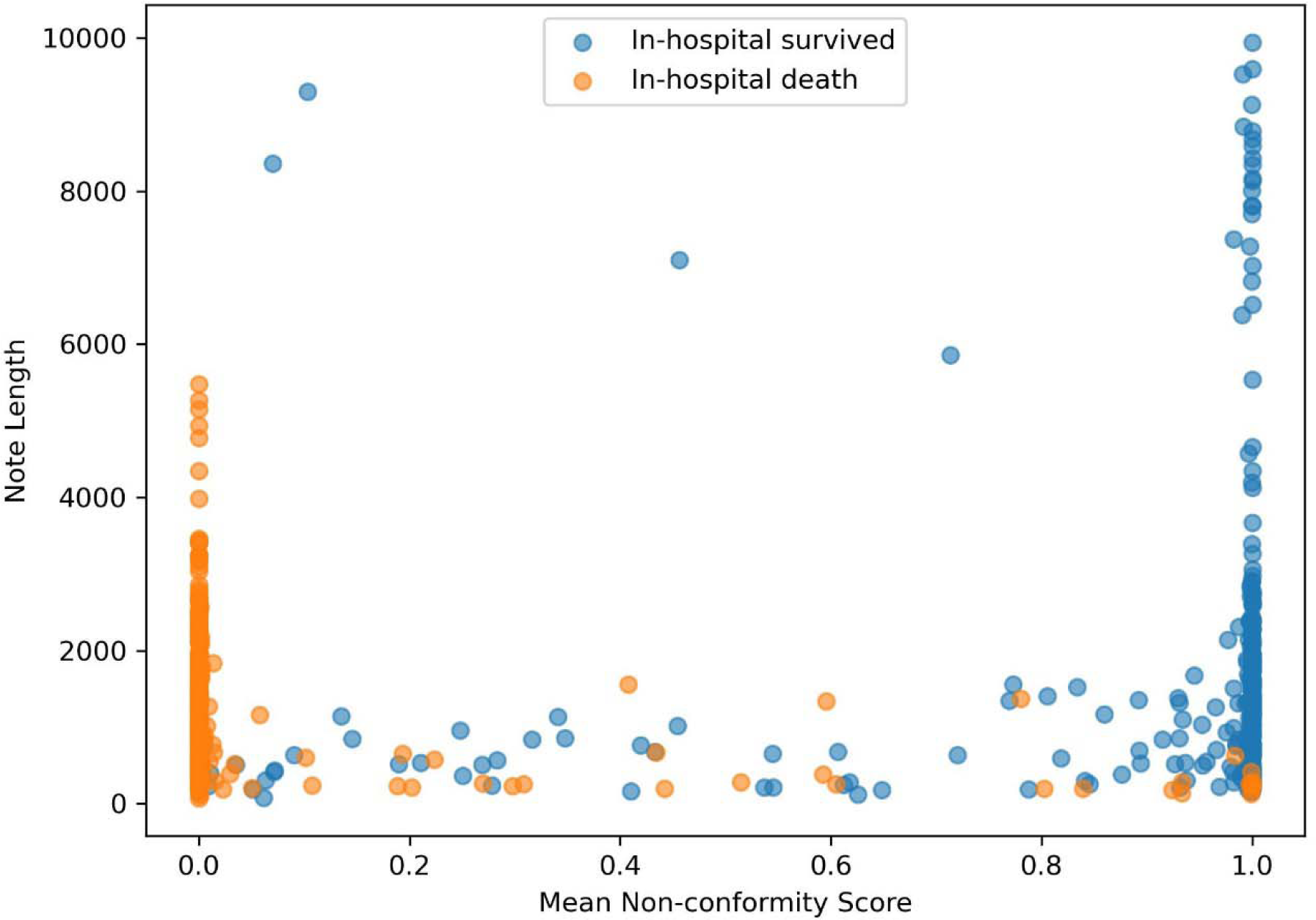
Scatter plot of nonconformity scores versus note length, stratified by in-hospital death outcome. Each point represents a patient from the calibration set. The y-axis shows the total number of tokens in the concatenated clinical note documented within the first 24 hours of admission. The x-axis represents the nonconformity score calculated for the true class label using conformal prediction.

**Table 1:**
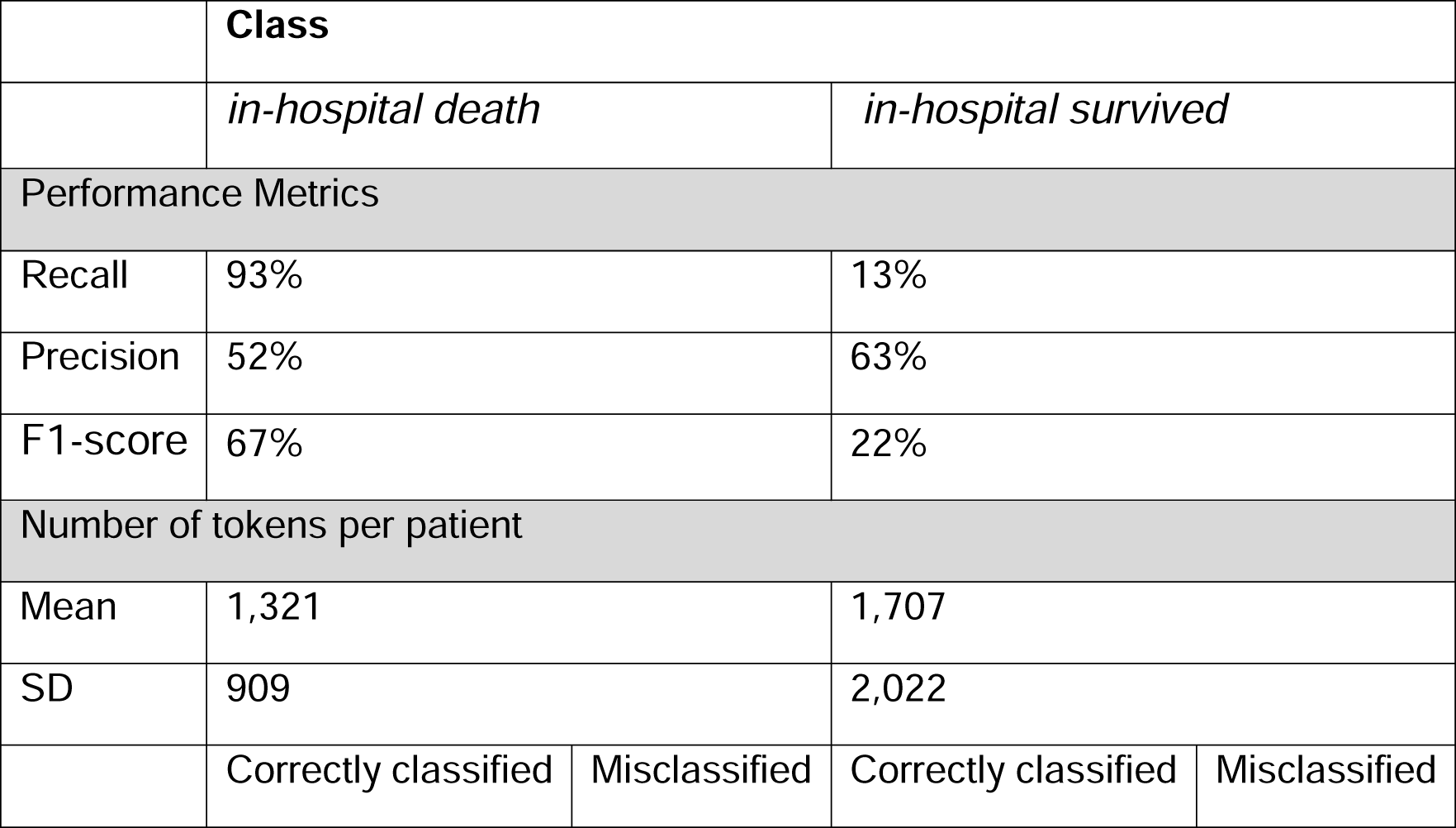
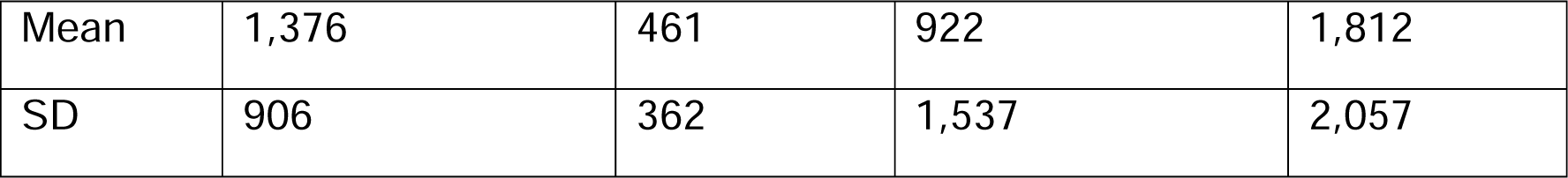
Note Length and LLM-Only Prediction Performance by Death Status.

On a balanced test set, the conformal prediction framework achieved an overall empirical coverage of 90.4%, slightly above the nominal 90% level, indicating conservative calibration. Disaggregated by class, coverage was 99.7% for “in-hospital death” (over-coverage) and 81.1% for *“*in-hospital survive” (under-coverage). The mean prediction set size was 1.67 labels overall, with “in-hospital survive” instances producing larger sets (mean = 1.80) than “in-hospital death” instances (mean 1.54), reflecting greater uncertainty for survival cases.

These results reveal a pronounced class-dependent calibration disparity. The underlying classifier assigns consistently higher predictive probabilities—and thus higher p-values—to the “in-hospital death” class, causing most positive instances to exceed the global p-value threshold and achieve near-perfect coverage (99.7%). Conversely, the model expresses lower confidence in the “in-hospital survive” class, resulting in more frequent exclusion from prediction sets, undercoverage (81.1%), and larger average prediction set sizes.

This imbalance is further evident in the distribution of prediction set sizes. Among all conformal predictions, 32.9% produced singleton sets (representing confident predictions where the model assigned only one class), while 67.1% were hedged cases containing multiple possible outcomes, indicating calibrated uncertainty in borderline scenarios [58]. The class-specific breakdown reinforces this pattern: “in-hospital survive” cases yielded only 20.1% singleton sets (79.9% multiple outcome), whereas “in-hospital death” cases produced 45.8% singleton sets (54.2% multiple outcome), with no empty sets observed in either class. This distribution reveals a clear class-specific stratification of uncertainty, with survival predictions exhibiting systematically higher uncertainty than mortality predictions.

Importantly, this class-conditional calibration imbalance represents a known limitation of the standard split-conformal prediction framework, which enforces marginal coverage over the entire test set but does not guarantee class-specific calibration. As a result, when the learned p-value threshold is even slightly too permissive, the method tends to favor overcoverage overall, while still allowing class-level imbalances to persist.

These findings suggest that class-conditional conformal prediction methods may be necessary in contexts where balanced coverage across classes is critical— particularly when the clinically relevant outcome is a minority class—though such approaches reduce effective calibration set size and warrant careful consideration of this tradeoff when class imbalance is moderate or when the outcome currently favored by the model aligns with clinical priorities.

Figure 4 displays the relationship between credibility (conformal p-value of the most likely class) and confidence (one minus the p-value of the second most likely class) in GPT predictions on the test set. The strong positive relationship indicates that high statistical support for the top prediction typically coincides with clear separation from competing alternatives. Most predictions are correctly covered (orange points along the upward trend). However, a small cluster of blue points at high credibility reveals a form of overconfidence: the model occasionally assigns high statistical support to incorrect predictions. In an ideally calibrated model, we expect errors to cluster around low credibility (low-confidence) regions, but this pattern indicates the model sometimes exhibits overconfidence on mistakes.

**Figure 4:**
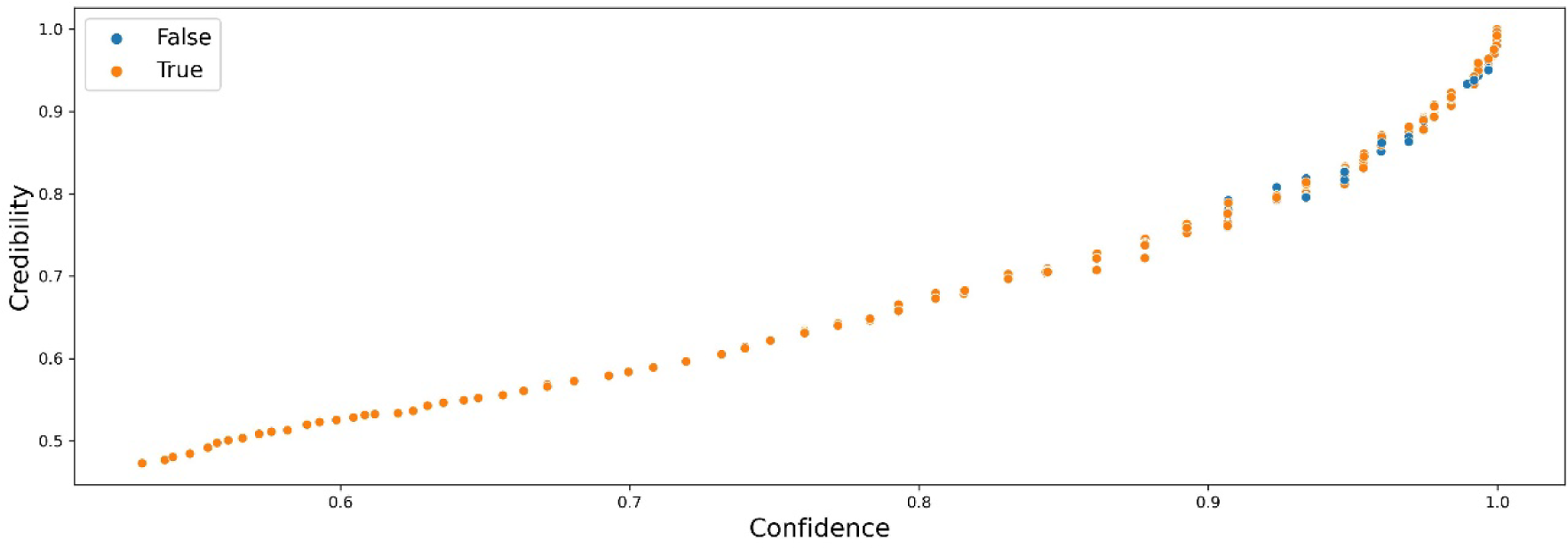
Credibility vs. Confidence in GPT Predictions. Each point is a model prediction; dots are color-coded by conformal coverage: orange = covered (correct), blue = miscoverage (incorrect). Credibility reflects statistical support for the top label, while confidence reflects separation from competing labels. Points follow a strong upward trend: higher credibility generally coincides with higher confidence, and most predictions are correctly covered (orange). Notably, a small cluster of blue points at high credibility indicates overconfident errors: cases where the model assigns high support to an ultimately incorrect label.

## Discussion

The GPT performed substantially better in predicting death cases than survival. It correctly identified around 94% of the patients who died in the hospital, indicating a strong sensitivity or recall for the positive class (“in-hospital death”). In contrast, it correctly predicted survival in only around 22% of the cases. Prioritizing sensitivity for in-hospital death can be clinically advantageous in high-stakes settings where the consequence of missed deterioration is severe. A model that preferentially captures in-hospital death cases can serve as an early warning layer to trigger rapid evaluation, closer monitoring, and timely escalation of care. In practice, this bias can help surface a manageable subset of patients for proactive interventions, support dynamic resource allocation, and focus team attention on those at greatest imminent risk. Moreover, this imbalance suggests that the model tends to overpredict death, possibly due to characteristics in the notes that bias the model towards more critical outcomes, or due to limitations in distinguishing subtle signals of recovery from high-risk notes.

The relationship observed between note length and nonconformity scores aligns closely with the patterns reported in Table 1. Patients who died during hospitalization had slightly shorter notes on average compared to those who survived. However, when stratified by prediction outcome, a clearer pattern emerges: for death cases, longer notes were associated with correct classification, whereas for survival cases, longer, heterogeneous documentation more often coincided with misclassification. This is supported by the lower nonconformity scores for most death cases in Figure 2, indicating high model confidence when sufficient clinical detail is present.

The behavior observed in Figure 4 signals that conformal prediction may not effectively compensate for overconfident but incorrect model outputs. While most predictions show a consistent relationship between high confidence and accurate outcomes, this figure reveal a recurring failure mode, overconfident but incorrect predictions, even in the presence of calibration. Relying solely on GPT’s internal probability estimates even when augmented with conformal prediction, could therefore mislead clinicians if model failures are masked by overconfidence. These findings underscore the need for finer-grained calibration approaches, potentially incorporating class-conditional or adaptive conformal techniques, to more effectively mitigate overconfident errors and improve the reliability of AI-assisted decision-making.

This study fills several important gaps in the existing literature on clinical risk prediction. Prior research has largely focused on structured EHR variables, such as laboratory values, vitals, or demographic data, which, while valuable, capture only a fraction of the contextual and temporal nuance present in narrative documentation. By leveraging GPT-4o to interpret early documented ICU notes, this work investigates the applicability of LLMs for early prediction in critically ill patients, and finds that, without task-specific fine-tuning, their discriminative performance is limited. Furthermore, by integrating conformal prediction, we move beyond the traditional performance-centric focus of prior models to explicitly quantify and calibrate patient-level uncertainty, an area rarely addressed in prior LLM-based clinical prediction studies.

### Conclusion and future work

This work investigates the prediction capability of GPT based solely on clinical notes documented within the first 24 hours of ICU patient admission and integrates conformal prediction to quantify and calibrate the uncertainty of model outputs. Through the integration of conformal prediction, we quantified uncertainty in model outputs and achieved empirical coverage exceeding the nominal 90% level. However, deeper analysis of credibility, confidence, and nonconformity scores revealed critical limitations. Specifically, the model exhibited overconfidence in certain incorrect predictions, leading to notable undercoverage in survival cases and highlighting a class-conditional calibration imbalance. These findings underscore the importance of uncertainty-aware modeling in clinical decision support. Future work should explore adaptive or class-conditional conformal prediction frameworks, fine-tune language models using domain-specific data, and evaluate hybrid models that integrate structured EHR features with unstructured text to achieve more balanced and reliable predictions. Additionally, our reliance on the MIMIC-III dataset, though comprehensive, restricts the generalizability of our findings. The dataset’s inherent biases and its derivation from a specific clinical environment may not accurately reflect the varied patient populations found across different geographic or healthcare settings. Despite these limitations, the study presents a significant step forward in our understanding of the capabilities and limitations of advanced language models in the critical domain of healthcare.

## Ethics Approval

No protected health information was collected, and the analytical dataset was fully de-identified.

## Data Availability

The reader can reproduce our experiments by gaining access to the MIMIC-III dataset and using our public https://doi.org/10.5281/zenodo.15793500

## Acknowledgements

The research reported in this publication was supported in part by the National Center for Advancing Translational Sciences of the National Institutes of Health under Award Number UM1TR004409 . This work was supported by the Department of Defense (DoD) SMART Scholarship-for-Service Program to AM. The content is solely the responsibility of the authors and does not necessarily represent the official views of the Department of Defense or the National Institutes of Health.

## Contributorship Statement

Fatemeh Shah-Mohammadi: Conceptualization, Data curation, Formal analysis, Investigation, Methodology, Validation, Visualization, Writing – original draft, Writing – review & editing. Alexander Millar: Conceptualization, Data curation, Formal analysis, Methodology, Validation, Visualization, Writing – original draft, Writing – review & editing. Julio Facelli: Conceptualization, Funding acquisition, Methodology, Supervision, Writing – review & editing. Ram Gouripeddi: Conceptualization, Methodology, Supervision, Writing – review &editing.

## References

1. Lee S, Hong S, Cha WC, et al. (2020). Predicting adverse outcomes for febrile patients in the emergency department using sparse laboratory data: development of a time adaptive model. JMIR Medical Informatics, 8, e16117.

2. Hong S, Lee S, Lee J, et al. (2020). Prediction of cardiac arrest in the emergency department based on machine learning and sequential characteristics: model development and retrospective clinical validation study. JMIR Medical Informatics, 8, e15932.

3. Kwon JM, Lee Y, Lee Y, et al. (2018). Validation of deep-learning-based triage and acuity score using a large national dataset. PLoS ONE, 13, e0205836.

4. Redfern O C, Pimentel M A F, Prytherch D, et al. (2018). Predicting in-hospital mortality and unanticipated admissions to the intensive care unit using routinely collected blood tests and vital signs: Development and validation of a multivariable model. Resuscitation, 133, 75–81.

5. Silva I, Moody G, Scott DJ, et al. (2012). Predicting in-hospital mortality of ICU patients: The PhysioNet/Computing in Cardiology Challenge 2012. Computing in Cardiology, IEEE, 245–248.

6. Fernandes M, Vieira SM, Leite F, et al. (2020). Clinical decision support systems for triage in the emergency department using intelligent systems: a review. Artificial Intelligence in Medicine, 102.

7. Ghassemi M, Celi LA, & Stone DJ. (2015). State of the art review: the data revolution in critical care. Critical Care, 19(1), 1–9.

8. Gilboy N, Tanabe T, Travers D, et al. (2011). Emergency Severity Index (ESI): A triage tool for emergency department. Agency for Healthcare Research and Quality, Rockville, MD.

9. Knaus WA. (2002). APACHE 1978–2001: the development of a quality assurance system based on prognosis: milestones and personal reflections. Archives of Surgery, 137(1), 37– 41.

10. Lemeshow S, Teres D, Klar J, et al. (1993). Mortality Probability Models (MPM II) based on an international cohort of intensive care unit patients. JAMA, 270(20), 2478–2486.

11. Toma T, Abu-Hanna A, & Bosman RJ. (2007). Discovery and inclusion of SOFA score episodes in mortality prediction. Journal of Biomedical Informatics, 40(6), 649–660.

12. Levin S, Toerper M, Hamrock E, et al. (2018). Machine-learning-based electronic triage more accurately differentiates patients with respect to clinical outcomes compared with the emergency severity index. Annals of Emergency Medicine, 71(5), 565–574.e2.

13. Keegan M T, Gajic O, & Afessa B. (2011). Severity of illness scoring systems in the intensive care unit. Critical Care Medicine, 39(1), 163–169.

14. Johnson AEW, Pollard TJ, Shen L, et al. (2016). MIMIC-III, a freely accessible critical care database. Scientific Data, 3(1), 1–9. 10.1038/sdata.2016.35.

15. Hyland SL, Faltys M, Hüser, M, Lyu X, et al. (2020). Early prediction of circulatory failure in the intensive care unit using machine learning. Nature Medicine, 26(3), 364–373. 10.1038/s41591-020-0789-4.

16. Goldberger A, Amaral LAN, Glass L, et al. (2000). PhysioBank, PhysioToolkit, and PhysioNet: Components of a new research resource for complex physiologic signals. Circulation, 101(23), e215–e220.

17. Lee M, et al. (2020). WiDS (Women in Data Science) Datathon 2020: ICU Mortality Prediction (version 1.0.0). PhysioNet. 10.13026/vc0e-th79.

18. Minaee S, Mikolov T, Nikzad N, et al. (2024). Large Language Models: A Survey. arXiv. 10.48550/arXiv.2402.06196.

19 Zhou J, Li T, Fong SJ, Dey N, Crespo RG. Exploring chatGPT’S potential for consultation, recommendations and report diagnosis: Gastric cancer and gastroscopy reports’ case. IJIMAI. 2023;8(2):7–13.

20 Choi HS, Song JY, Shin KH, Chang JH, Jang BS. Developing prompts from large language model for extracting clinical information from pathology and ultrasound reports in breast cancer. Radiation Oncology Journal. 2023;41(3):209.

21. Singhal K, Azizi S, Tu T, et al. Large language models encode clinical knowledge. arXiv preprint arXiv:2212.13138. 2022 Dec 26.

22. Ouyang L, Wu J, Jiang X, Almeida D, et al. Training language models to follow instructions with human feedback. Advances in Neural Information Processing Systems. 2022 Dec 6;35:27730–44.

23. EMR vs. EHR: Understand the Difference | Elevance Health. www.Elevancehealth.Com.

24. Miller DD, Brown EW. Artificial Intelligence in Medical Practice: The Question to the Answer? Am J Med 2018;131:129–33. 10.1016/j.amjmed.2017.10.035.

25. Marr B. Revolutionizing Healthcare: The Top 14 Uses Of ChatGPT In Medicine And Wellness. Forbes n.d. https://www.forbes.com/sites/bernardmarr/2023/03/02/revolutionizing-healthcare-the-top-14-uses-ofchatgpt-in-medicine-and-wellness/ (accessed June 29, 2023).

26. Rao A, Kim J, Kamineni M, Pang M, et al. Evaluating ChatGPT as an Adjunct for Radiologic Decision-Making. MedRxiv Prepr Serv Health Sci 2023:2023.02.02.23285399. 10.1101/2023.02.02.23285399.

27. Shen Y, Heacock L, Elias J, et al. ChatGPT and Other Large Language Models Are Double-edged Swords. Radiology 2023;307:e230163. 10.1148/radiol.230163.

28. About us. SNOMED Int n.d. https://www.snomed.org/about-us (accessed July 5, 2023).

29. Moons P, Van Bulck L. ChatGPT: can artificial intelligence language models be of value for cardiovascular nurses and allied health professionals. Eur J Cardiovasc Nurs 2023:zvad022. 10.1093/eurjcn/zvad022.

30. Xie Q, Wang F. Faithful AI in Healthcare and Medicine. MedRxiv 2023. 10.1101/2023.04.18.23288752.

31. Liu S, Wright AP, Patterson BL, et al. Assessing the Value of ChatGPT for Clinical Decision Support Optimization. MedRxiv Prepr Serv Health Sci 2023:2023.02.21.23286254. 10.1101/2023.02.21.23286254.

32 Rosen S, Saban M. Evaluating the reliability of ChatGPT as a tool for imaging test referral: a comparative study with a clinical decision support system. European Radiology. 2024 May;34(5):2826–37.

33 Wang S, Zhao Z, Ouyang X, Wang Q, Shen D. ChatCAD: Interactive Computer-Aided Diagnosis on Medical Image using Large Language Models. ArXivOrg 2023. 56. Harrer S. Attention is not all you need: the complicated case of ethically using large language models in healthcare and medicine. EBioMedicine 2023;90:104512. 10.1016/j.ebiom.2023.104512.

34 Y LeCun. 2023. Do large language models need sensory grounding for meaning and understanding? In Workshop on Philosophy of Deep Learning, NYU Center for Mind, Brain, and Consciousness and the Columbia Center for Science and Society.

35 Vladimir Vovk, Alexander Gammerman, and Glenn Shafer. 2005. Algorithmic learning in a random world, volume 29. Springer.

36. Angelopoulos AN, Bates S. A gentle introduction to conformal prediction and distribution-free uncertainty quantification. arXiv preprint arXiv:2107.07511. 2021 Jul 15.

37 Yuko Kato, David MJ Tax, and Marco Loog. 2023. A review of nonconformity measures for conformal pre-diction in regression. Conformal and Probabilistic Prediction with Applications, pages 369–383.

38 Mauricio Sadinle, Jing Lei, and Larry Wasserman. 2019. Least ambiguous set-valued classifiers with bounded error levels. Journal of the American Statistical Association, 114(525):223–234.

39 Bhawesh Kumar, Charlie Lu, Gauri Gupta, Anil Palepu, David Bellamy, Ramesh Raskar, and Andrew Beam. 2023. Conformal prediction with large language models for multi-choice question answering. arXiv preprint arXiv:2305.18404.

40 Anastasios N Angelopoulos, Stephen Bates, Emmanuel J Candès, Michael I Jordan, and Lihua Lei. 2021. Learn then test: Calibrating predictive algorithms to achieve risk control. arXiv preprint arXiv:2110.01052.

41 Victor Quach, Adam Fisch, Tal Schuster, Adam Yala, Jae Ho Sohn, Tommi S Jaakkola, and Regina Barzilay. 2023. Conformal language modeling. arXiv preprint arXiv:2306.10193.

42 Khanh Nguyen and Brendan O’Connor. 2015. Posterior calibration and exploratory analysis for natural language processing models. arXiv preprint arXiv:1508.05154.

43 Stephanie Lin, Jacob Hilton, and Owain Evans. 2022. Teaching models to express their uncertainty in words. arXiv preprint arXiv:2205.14334.

44 Su J, Luo J, Wang H, Cheng L. Api is enough: Conformal prediction for large language models without logit-access. arXiv preprint arXiv:2403.01216. 2024 Mar 2.

45 Lee S, Lee HH, Lee H, Yum KS, Baek JH, Khil J, Lee J, Shin S, Cho M, Ahn NY, You SC. Confidence-linked and uncertainty-based staged framework for phenotype validation using large language models. Journal of the American Medical Informatics Association. 2025 Jun 17:ocaf099.

46 Achiam J, Adler S, Agarwal S, Ahmad L, Akkaya I, Aleman FL, Almeida D, Altenschmidt J, Altman S, Anadkat S, Avila R. Gpt-4 technical report. arXiv preprint arXiv:2303.08774. 2023 Mar 15

47 Tripathi S, Alkhulaifat D, Doo FX, Rajpurkar P, McBeth R, Daye D, Cook TS. Development, Evaluation, and Assessment of Large Language Models (DEAL) Checklist: A Technical Report. NEJM AI. 2025 May 22;2(6):AIp2401106.

48 Xiao C, Choi E, Sun J. Opportunities and challenges in developing deep learning models using electronic health records data: a systematic review. J Am Med Inform Assoc. 2018 Oct 1;25(10):1419–1428. doi: 10.1093/jamia/ocy068. PMID: 29893864; PMCID: PMC6188527.

49 Van Calster, B., McLernon, D.J., van Smeden, M. et al. Calibration: the Achilles heel of predictive analytics. BMC Med 17, 230 (2019). 10.1186/s12916-019-1466-7

50 Yang, X., Chen, A., PourNejatian, N. et al. A large language model for electronic health records. npj Digit. Med. 5, 194 (2022). 10.1038/s41746-022-00742-2

51 Weissman GE, Hubbard RA, Ungar LH, Harhay MO, Greene CS, Himes BE, Halpern SD. Inclusion of Unstructured Clinical Text Improves Early Prediction of Death or Prolonged ICU Stay. Crit Care Med. 2018 Jul;46(7):1125–1132. doi: 10.1097/CCM.0000000000003148. PMID: 29629986; PMCID: PMC6005735.

52 Mahbub M, Srinivasan S, Danciu I, Peluso A, Begoli E, Tamang S, Peterson GD. Unstructured clinical notes within the 24 hours since admission predict short, mid & long-term mortality in adult ICU patients. PLoS One. 2022 Jan 6;17(1):e0262182. doi: 10.1371/journal.pone.0262182. PMID: 34990485; PMCID: PMC8735614.

53 Brown T, Mann B, Ryder N, Subbiah M, Kaplan JD, Dhariwal P, Neelakantan A, Shyam P, Sastry G, Askell A, Agarwal S. Language models are few-shot learners. Advances in neural information processing systems. 2020;33:1877–901.

54 Rajkomar, A., Oren, E., Chen, K. et al. Scalable and accurate deep learning with electronic health records. npj Digital Med 1, 18 (2018). 10.1038/s41746-018-0029-1.

55 Begoli, E., Bhattacharya, T. & Kusnezov, D. The need for uncertainty quantification in machine-assisted medical decision making. Nat Mach Intell 1, 20–23 (2019). 10.1038/s42256-018-0004-1

56 Williams CYK, Zack T, Miao BY, Sushil M, Wang M, Kornblith AE, Butte AJ. Use of a Large Language Model to Assess Clinical Acuity of Adults in the Emergency Department. JAMA Netw Open. 2024 May 1;7(5):e248895. doi: 10.1001/jamanetworkopen.2024.8895. PMID: 38713466; PMCID: PMC11077390.

57 Sarbay İ, Berikol GB, Özturan İU. Performance of emergency triage prediction of an open access natural language processing based chatbot application (ChatGPT): A preliminary, scenario-based cross-sectional study. Turk J Emerg Med. 2023 Jun 26;23(3):156–161. doi: 10.4103/tjem.tjem_79_23. PMID: 37529789; PMCID: PMC10389099.

58 Bellotti A, Zhao X. Conformal Prediction and Trustworthy AI. arXiv.org. Published 2025. Accessed October 21, 2025. https://arxiv.org/abs/2508.06885.

