## Appendix 1 for "Understanding Uncertainty in Large Language Model Predictions of Early Death in Critically Ill Patients: A Conformal Prediction Approach"

The following lists the intermediate prompts that are iteratively evaluated for clarity and consistency of response formatting:

Prompt 1:

“Read the following clinical note and answer: Will the patient die during this hospital admission? <clinical note text here>”

This prompt was too open-ended and lacked clear constraints, which often led GPT to generate full-sentence explanations or provide contextual reasoning rather than a concise binary answer.

Prompt 2:

“Based on the following clinical note from the first day of admission, predict whether the patient is at risk of dying during this hospital stay. Respond with either “Yes” or “No” only. <clinical note text here>”

While better than Prompt 1, GPT still sometimes ignored the instruction to return just “Yes” or “No”, especially when the clinical note was too long.

Prompt 3:

“You are a clinical decision support assistant. Use the following clinical note from the first day of admission to decide if the patient is at risk of dying during this hospitalization. Answer only “Yes” or “No”. <clinical note text here>"

This prompt lacked an explicit call to analyze **specific clinical signals** (like vitals, labs, etc.), which could reduce focus and consistency across notes of varying quality.
