## Appendix 2 for "Understanding Uncertainty in Large Language Model Predictions of Early Death in Critically Ill Patients: A Conformal Prediction Approach"

Table 1 presents the distribution of in-hospital death across ethnicity, gender, and marital status. Among ethnic groups, White patients constituted the largest proportion, with 37.65% of survivors and 38.26% of those who died. Black patients accounted for 8.16% of survivors and 4.80% of deaths, while Hispanic patients made up 1.60% of survivors and 0.94% of deceased cases. Asian patients represented 1.38% of survivors and 1.21% of deaths. Patients categorized under "Other" ethnicity comprised 2.05% of survivors and a notably higher 3.97% of deaths. Regarding gender, female patients made up 22% of survivors and 23% of deaths, whereas male patients constituted 29% of survivors and 27% of deaths. For marital status, 22.27% of survivors and 22% of deaths were married, 3.64% of survivors and 2.09% of deaths were divorced, 9.15% of survivors and 11.69% of deaths were widowed, and patients categorized as "Other" made up 2.15% of survivors and 4.63% of deaths.

Table 1: Demographic Statistics.

|  | **Death** | **Percentage (%)** |
| --- | --- | --- |
| **Ethnicity** | | |
| Other | No | 2.05 |
|  | Yes | 3.97 |
| Asian | No | 1.38 |
|  | Yes | 1.21 |
| Black | No | 8.16 |
|  | Yes | 4.80 |
| Hispanic | No | 1.60 |
|  | Yes | 0.94 |
| White | No | 37.65 |
|  | Yes | 38.26 |
| **Gender** | | |
| Female | No | 22.00 |
|  | Yes | 23.00 |
| Male | No | 29.00 |
|  | Yes | 27.00 |
| **Marital Status** |  |  |
| Divorced | No | 3.64 |
|  | Yes | 2.09 |
| Married | No | 22.27 |
|  | Yes | 22.00 |
| Other (Single + Unknown) | No | 2.15 |
|  | Yes | 4.63 |
| Widowed | No | 9.15 |
|  | Yes | 11.69 |
